# Attained body mass index among children attending outdoor or conventional kindergartens

**DOI:** 10.1101/2022.10.03.22280495

**Authors:** Sofus C. Larsen, Jeanett F. Rohde, Nanna J. Olsen, Jane N. Østergaard, Berit L. Heitmann, Ina O. Specht

## Abstract

**BACKGROUND:** Studies have shown that outdoor activities are inversely associated with adiposity, but little is known about outdoor kindergartens as a potential intervention for healthy weight development.

**METHODS:** We included 1544 children from outdoor kindergartens and 1640 from conventional kindergartens. Anthropometry was measured by school health nurses when the children were 6 to 8 years old. The first available measure of attained body mass index z-score (BMIz) after school entrance was included as the primary outcome. Risk of attaining overweight (including obesity) was included as a secondary outcome. Register-based information was available on potential confounding factors. Regression models were constructed to examine differences in study outcomes between children attending outdoor or conventional kindergartens.

**RESULTS:** Our basic models, with information on outcome, kindergarten type and birth weight, showed a borderline statistically significantly lower attained BMIz (−0.07 [95% CI: −0.14, 0.00], P=0.060) and a lower risk of overweight (adjusted risk ratio: 0.83 [95% CI: 0.72, 0.97], P=0.016) among children attending outdoor kindergartens compared to conventional kindergartens. When adjusting for sociodemographic factors and parental BMI, there was no evidence of differences in attained BMIz (P=0.153) or overweight (P=0.967).

**CONCLUSION:** Our results do not support outdoor kindergartens as an intervention for healthy weight development.

**Study importance questions:** *What is already known about this subject?:*

- One of the many suggested explanations for the increased prevalence of childhood obesity is a decline in children’s outdoor activities.
- Previous studies have shown outdoor activities to be inversely associated with adiposity.
- Little is known about outdoor kindergartens as a potential structural intervention for healthy weight development.

*What are the new findings in your manuscript?:*

- Our crude analysis suggested a lower risk of attained overweight after school entry among children in outdoor kindergartens than children in conventional kindergartens.
- After adjusting for potential confounding from parental BMI and sociodemographic factors, we found no evidence of differences in attained BMIz or overweight among children attending outdoor or conventional kindergartens.

*How might your results change the direction of research or the focus of clinical practice?:*

- Our results do not support outdoor kindergartens as a potential intervention for healthy weight development.
- These results highlight the importance of adjusting for potential confounding factors when examining the relationship between outdoor activities and childhood obesity.
- Given the observational design of the study, our results should be interpreted with caution, and randomized trials are warranted to confirm our findings.

**Article Summary:** This study examined whether children in outdoor kindergartens had a lower risk of overweight after school entrance compared to children in conventional kindergartens.

## BACKGROUND

Between 1975 and 2016, the global prevalence of obesity among children and adolescents has increased from 0.9% to 7.8% among boys and from 0.7% to 5.6% among girls [1]. This has raised serious public health concerns, as childhood obesity is associated with both short- and long-term health consequences, including negative self-evaluation, bullying, social stigma, poor academic performance, symptoms of depression or anxiety [2, 3], numerous physical comorbidities [4] and an increased risk of mortality both in early [5] and later adulthood [6]. Once childhood obesity is established it is difficult to reverse [7] and children with obesity often grow up to be adults with obesity [8], strengthening the case for early intervention. However, the effect of existing prevention interventions are modest at best [9], indicating that alternative approaches may be needed to effectively promote healthy weight development in childhood.

One of the many suggested explanations for the increased prevalence of childhood obesity is a decline in children’s outdoor activities [10]. In support of this, some cross-sectional studies have shown both outdoor play time and the quality of the outdoor environment to be inversely associated with childhood body mass index (BMI), waist circumference and risk of obesity [11-14]. While the evidence from longitudinal studies is limited, results based on 2810 children from the *Child Experiences Survey 2006* cohort suggested that outdoor play time was associated with decreases in children’s BMI over the course of a preschool year [12]. Likewise, Armstrong and colleagues (2015) showed a higher density of parks or other recreational locations to be associated with a decrease in BMI z-score (BMIz) over time among 93 children with overweight involved in a family-based weight management intervention [15]. However, even though many of the published studies suggest some beneficial effects of outdoor activities on adiposity among children, the evidence is far from consistent, as pointed out in a recent systematic review on the relationship between nature exposure and different measures of childhood health [11]. In addition, there is a need to develop and evaluate interventions that can be implemented at a larger scale and in a cost–effective manner.

An ideal setting for conducting childhood interventions is the kindergarten, where many children spend a large part of their daily lives and may establish some of the habits and health behaviors that can track into later childhood and adulthood [16]. Larger cities in Denmark and the other Scandinavian countries have two different types of kindergartens: 1) the conventional kindergarten were the children have access to playgrounds but also spend some of their day indoor, and 2) the outdoor kindergarten where most or all of the day is spent playing outside, often in a forest setting environment [17]. In a pre-test-post-test study of 74 children with accelerometer measured information on physical activity, we have previously shown that children are more physically active in an outdoor kindergarten setting than in a conventional kindergarten setting [18]. However, the potential benefits of the Danish outdoor kindergartens on promotion of healthy weight development have not been thoroughly investigated.

Combined with the unique Danish health registers and systematic measurements of height and weight performed by school health nurses, we had a unique opportunity to examine outdoor kindergartens as a potential intervention setting for preventing development of overweight later in childhood. Our primary objective was to examine whether children in outdoor kindergartens had attained a lower BMIz and risk of overweight (including obesity) after school entrance compared to children in conventional kindergartens, based on the first available measure of outcome recorded by school health nurses when children were aged between 6 and 8 years. As secondary objectives, we examined potential effect modification by socioeconomic status, and the trajectories of attained BMIz and risk of overweight at 6, 7 and 8 years of age for children in outdoor kindergartens and conventional kindergartens.

## MATERIALS AND METHODS

### Study design

The present study was part of the *‘Outdoor kindergartens - the healthier choice?’* (ODIN) project, where data was obtained from conventional kindergartens with many indoor activities, and outdoor kindergartens where most or all of the daily activities took place outside [17]. We had information on a total of 5077 children from the two largest municipalities in Denmark (Aarhus and Copenhagen), comprising all children who attended outdoor kindergartens (n=2434) and a sub-set of children who attended conventional kindergartens (n=2643). Data from outdoor and conventional kindergartens was obtained from the same areas of parental residence. In the primary analysis, we excluded all participants with missing data on one or more covariates (n=729) or follow-up information on outcome (n=1164), resulting in a total of 3184 children for the complete case analysis. In total, 1544 of these children were from outdoor kindergartens and 1640 were from conventional kindergartens. A flowchart illustrating inclusion/exclusion of individuals can be found in the supporting information (**Figure S1**).

### Ethical considerations

Permission from the two municipalities to send information to Statistics Denmark was granted. Ethical permission was decided not to be relevant by the Ethical Committee of the Capital Region of Denmark (journal nr.: H-19053587). Permission from the Capital Region Data Agency and the Danish Patient Safety Authority was granted (Journal nr.: P-2020-54 and 31-1521-8, respectively).

### Outcomes

Height and weight were measured by school health nurses when the children were 6 to 8 years old. BMI was calculated as weight in kilograms per height in meters squared. We then generated BMIz using a power transformation in increments of 0.1 years, applying national reference scores to the study population [19]. The first available measure of attained BMIz was included as our primary outcome, while the first available measure of attained overweight (including obesity) (defined as a BMIz>1 [yes/no]) was included as a secondary outcome.

### Covariates

Gender of the child (boys/girls), birth weight (g), preterm birth (yes/no), maternal age (years), maternal pre-pregnancy BMI (kg/m^2^) and information on maternal smoking during pregnancy (yes/no) was obtained from the *Danish Medical Birth Register*. Information on maternal education was obtained from *Statistics Denmark* and included in the present study as *basic* (basic school 8th– 10th class), *short* (general upper–secondary education, short-cycle higher education or vocational education and training), *medium* (medium-cycle higher education or bachelor), and *long* (long-cycle education and PhD). Information on maternal country of origin (Western/non-Western) was also obtained from *Statistics Denmark*. Age at kindergarten enrolment (years) and total time spent in kindergarten (years) was obtained from the municipalities.

#### Additional covariates only included in sensitivity analyses

Information on paternal education and country of origin was obtained from *Statistics Denmark* and included the same categories as for maternal education and country of origin. In addition to birth weight, a subset of 2681 children had weight and length measured during the first year of life by health nurses at the home of the child. From this information, we included the last available measure of BMIz prior to kindergarten enrolment.

### Statistical analysis

A detailed statistical analysis plan, including estimates of power and precision, was circulated with all authors prior to conducting any statistical analyses **(Text S1)**.

Linear regression models were constructed to examine the mean difference in BMIz between children in outdoor kindergartens and conventional kindergartens based on the first recorded measure after school entry. We constructed a basic model with information on BMIz, kindergarten type and birth weight, and a fully adjusted model with added information on maternal age, maternal education, maternal country of origin, maternal smoking during pregnancy, maternal pre-pregnancy BMI, preterm birth, the child’s age at kindergarten enrolment, and total number of years spent in kindergarten.

Using the same adjustment strategy, we constructed logistic regression models to examine the risk of attaining overweight among children in outdoor kindergartens compared to conventional kindergartens based on the first recorded measure after school entry. We used the STATA postestimation command *adjrr* to attain adjusted estimates of absolute risks in addition to adjusted risk differences (aRD) and adjusted risk ratios (aRR) with corresponding 95% confidence intervals (95% CI) [20].

#### Between group differences in subgroups with low or high levels of maternal education

Low socioeconomic status is a strong predictor of childhood obesity [21] and previous results from the ODIN project have shown parental education to be higher for children attending outdoor kindergartens [22]. Thus, in addition to adjustment for maternal education, we conducted subgroup analyses specifically for children of mothers with a low level of education (defined as mothers with a basic or short education) and a high level of education (defined as mothers with a medium or long education). Effect modification by maternal education level was tested by adding the new 2-category education variable and a product term (kindergarten type × education) to the fully adjusted models after removing the original 4-category education variable.

#### Trajectories of attained BMIz and overweight

The same overall strategy was used to explore trajectories of attained BMIz and overweight at specific ages after school entry (6, 7 and 8 years of age). These analyses were restricted to children with at least one measurement of BMIz at 6 years of age. A last observation carried forward approach was then used for children with missing information at 7 and/or 8 years of age.

#### Sensitivity analyses

Since BMIz considers differential BMI trajectories by sex and age, we did not include sex as a potential confounder. However, effect modification of sex was tested by adding the variable *sex* and a product term (kindergarten type × sex) to the fully adjusted models.

We had no baseline measure of outcome available, but in addition to birth weight, most of the children had weight and length measured during the first year of life. From this, we added the last available measure of BMIz prior to kindergarten enrolment as a covariate.

We used maternal education as a proxy for the child’s SES. Similarly, we used maternal country of origin as a proxy for the geographical origin of the child. However, analyses were also conducted adding paternal information to the models.

We only included participants with complete information on all study variables, but as a sensitivity analysis multiple imputation by chained equations (MICE) was implemented in the complete sample of 5077 individuals.

Finally, we examined a potential dose–response relationship between time spent in kindergarten (continuous variable) and attained BMIz and tested for effect modification of kindergarten type by adding a product term to the model (years in kindergarten × kindergarten type).

All statistical tests were two-sided with a significance level at 0.05. All statistical analyses were performed using Stata/SE 16 (StataCorp LP, College Station, Texas, USA). Figures were produced using SigmaPlot 13.0 (San Jose, CA, USA).

## RESULTS

Characteristics of the included children can be found in **Table 1**. A total of 1544 children from outdoor kindergartens and 1640 from conventional kindergartens were included in the primary analysis. A lower proportion of girls were enrolled in outdoor kindergartens than in conventional kindergartens (43.9% vs 49.2%; P=0.003). Children in the outdoor kindergartens had a higher birth weight (3519 g [SD: 515] vs 3449 g [SD: 546]; P<0.001) and were less frequently born preterm (4.5% vs 7.1%, P=0.002). Statistically significant between-group differences were also observed among the children’s mothers, with a higher age (32.1 years [SD: 4.6] vs 31.4 [SD: 4.9]; P<0.001), lower pre-pregnancy BMI (22.4 [SD: 3.4] vs 23.1 [SD: 4.1]; P<0.001), higher proportion with a long education (46.2% vs 33.8%; P<0.001), lower proportion with a non-Western country of origin (3.6% vs 19.8%; P<0.001) and a lower proportion who smoked during pregnancy (6.3% vs 9.5% P=0.004) among mothers to children from outdoor kindergartens.

**Table 1.**
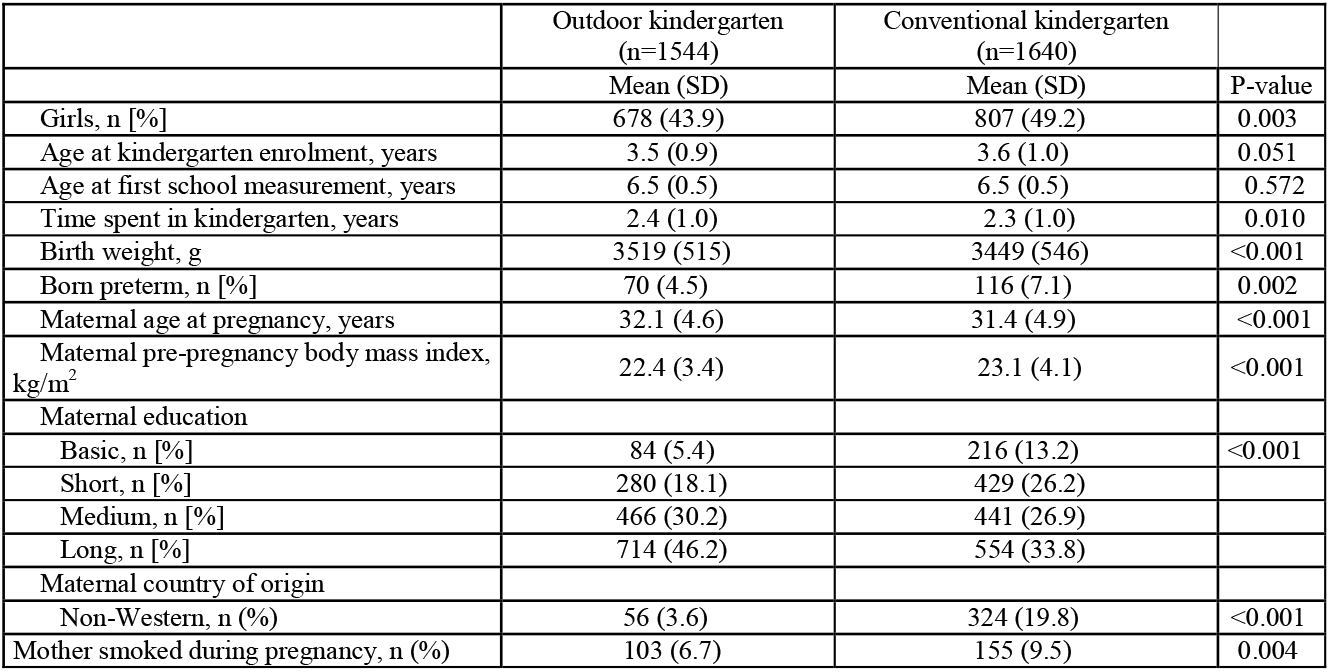
Participant characteristics of children in outdoor kindergartens versus conventional kindergartens. Results presented as mean (SD) unless otherwise stated.

The basic models with information on outcome, kindergarten type and birth weight, showed a borderline statistically significantly lower BMIz at school entry (−0.07 [95% CI: -0.14, 0.00], P=0.060) and lower risk of overweight (aRR: 0.83 [95% CI: 0.72, 0.97], P=0.016) among children in outdoor kindergartens compared to conventional kindergartens. However, in the full model with added information on sociodemographic factors we found no differences in BMIz (0.06 [95% CI: - 0.02, 0.12], P=0.153) or risk of overweight (aRR: 1.00 [95% CI: 0.86, 1.17], P=0.967) **(Table 2)**.

**Table 2.** Attained BMI z-scores and risk of overweight among children in outdoor kindergartens versus conventional kindergartens.

|  | Mean or % of participants (SE) |  | Estimated difference |  |
| --- | --- | --- | --- | --- |
|  | Outdoor kindergarten | Conventional kindergarten | Mean difference or aRD and aRR (95% CI) | P-value |
| **BMIz** |  |  |  |  |
| Basic model <sup>1</sup> | 0.05 (0.03) | 0.12 (0.03) | -0.07 (-0.14, 0.00) | 0.060 |
| Full model <sup>2</sup> | 0.11 (0.03) | 0.06 (0.02) | 0.06 (-0.02, 0.12) | 0.153 |
| **Overweight** |  |  |  |  |
| Basic model | 16.47% (0.93) | 19.77% (0.98) | aRD: - 3.29% (-5.96, 0.63)<br>aRR: 0.83 (0.72, 0.97) | 0.016<br>0.016 |
| Full model | 18.18% (0.99) | 18.13 (0.92) | aRD: 0.06% (-2.66, 2.77)<br>aRR: 1.00 (0.86, 1.17) | 0.967<br>0.967 |

Abbreviations: aRD, adjusted risk difference; aRR, adjusted risk ratio; BMIz, body mass index Z-score.
<sup>1</sup> Model with information on outcome, kindergarten type and birth weight.
<sup>2</sup> Added information maternal age, maternal education, maternal country of origin, maternal smoking during pregnancy, maternal pre-pregnancy body mass index, born preterm, age at kindergarten enrolment, and total number of years spent in kindergarten.

For both types of kindergartens, the highest BMIz after school entry was observed among children of mothers with a low level of education, but we found no effect modification by maternal education in relation to associations between kindergarten type and attained BMIz (P=0.512) or risk of overweight (P=0.154). In support of this, the fully adjusted models showed no between group differences in BMIz or risk of overweight in subgroups with mothers of low or high educational level (**Figure 1)**. Likewise, when comparing trajectories of attained BMI z-scores or overweight at 6, 7 and 8 years among children in outdoor kindergartens and conventional kindergartens, we did not find any statistically significant between-group differences (**Figure 2**).

**Figure 1.**
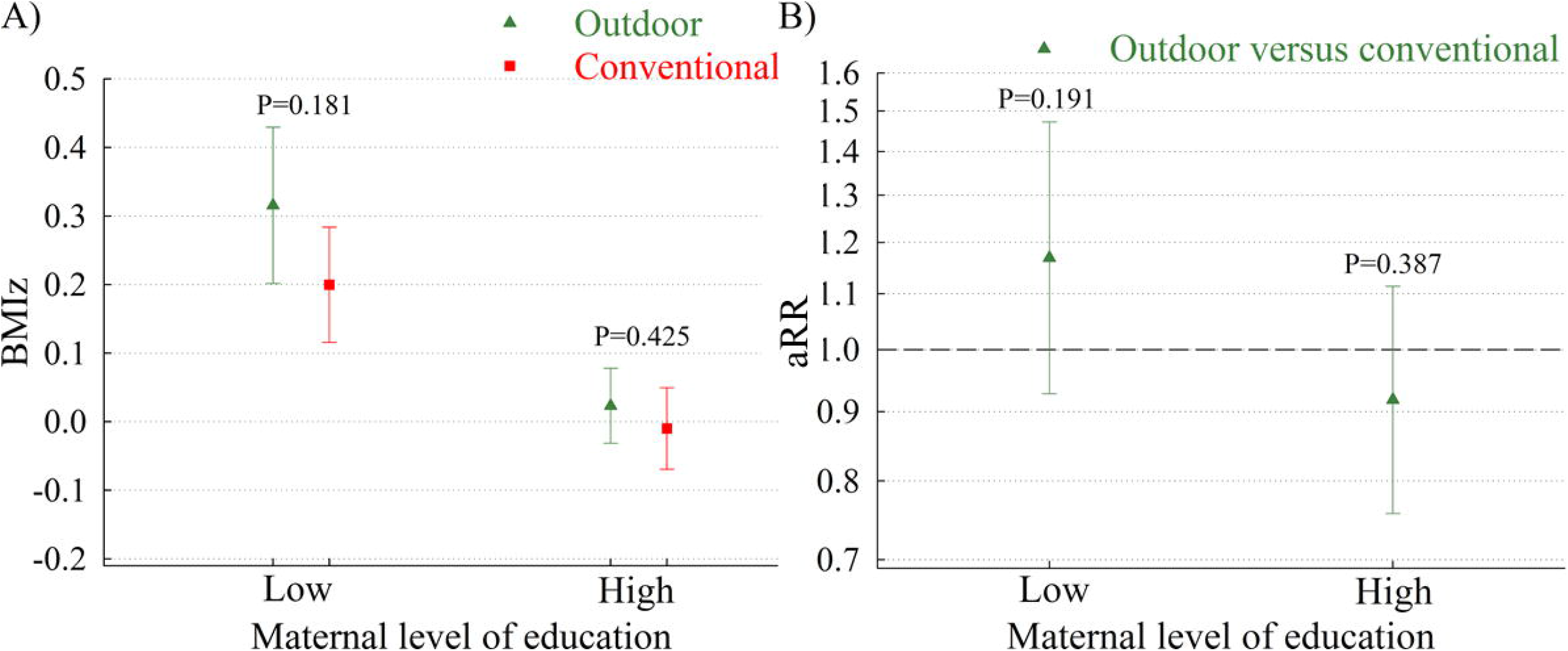
Attained BMI z-scores (A) and adjusted risk ratios for overweight (B) among children in outdoor kindergartens versus conventional kindergartens. Results stratified by maternal education. Abbreviations: aRR, adjusted risk ratios; BMIz, body mass index Z-score; SES, socioeconomic status. Low maternal education is defined as mothers with a basic or low education. High education is defined as mothers with a medium or high education. Results from model with information on outcome, kindergarten type, birth weight, maternal age, maternal country of origin, maternal smoking during pregnancy, maternal pre-pregnancy body mass index, preterm birth, age at kindergarten enrolment, and total number of years spent in kindergarten. Test statistics for BMIz: P-value for effect modification by education = 0.512. Test statistics for aRR: P-value for effect modification by education = 0.154.

**Figure 2.**
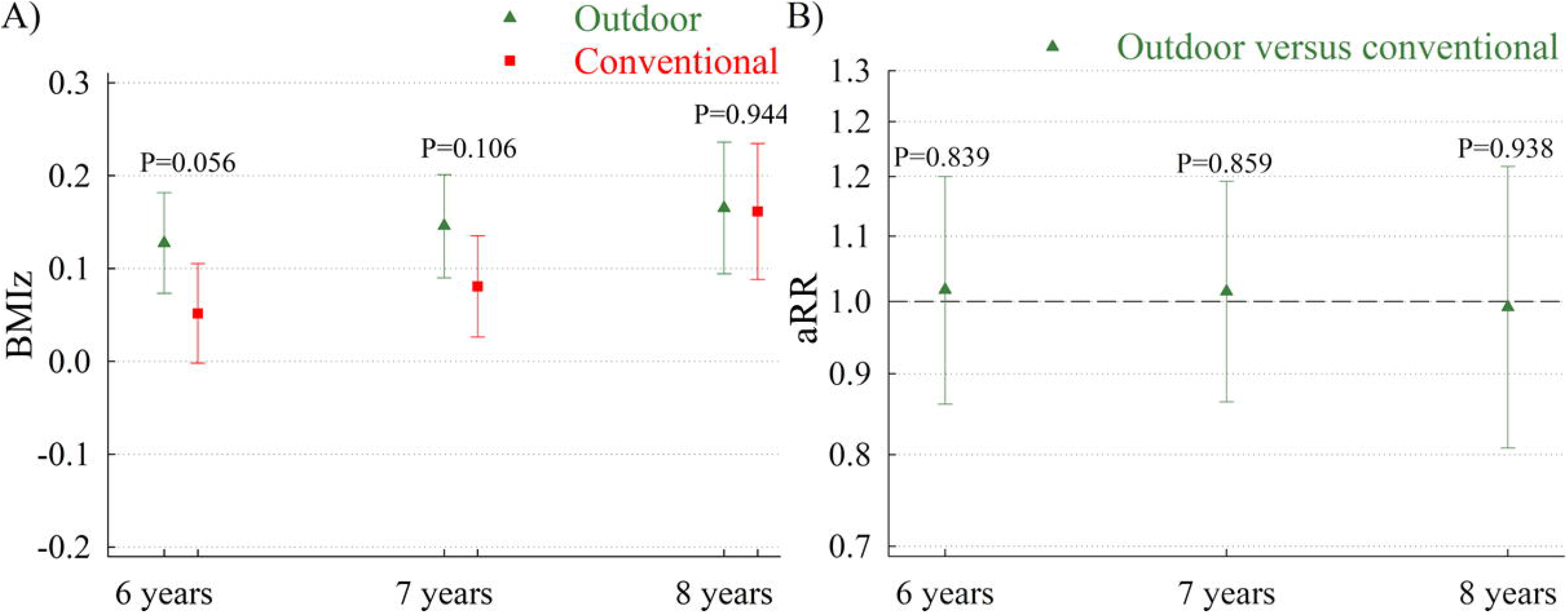
Trajectories of attained BMI z-scores (A) or overweight (B) at 6, 7 and 8 years of age among children in outdoor kindergartens versus conventional kindergartens. Abbreviations: BMIz, body mass index Z-score; aRR, adjusted risk ratios Results from model with information on outcome, kindergarten type, birth weight, maternal age, maternal country of origin, maternal smoking during pregnancy, maternal pre-pregnancy body mass index, preterm birth, age at kindergarten enrolment, and total number of years spent in kindergarten.

### Sensitivity analyses

We found no effect modification by sex in relation to BMIz (P=0.486) or aRR of obesity (P=0.800). After adding the last available measure of BMIz prior to kindergarten enrolment (**Table S1**) or paternal information on education and country of origin (**Table S2**) as covariates our results remained statistically non-significant. Likewise, analyses where missing information was handled through multiple imputation also remained statistically non-significant (**Table S3**). Finally, we found no association between years spend in outdoor or conventional kindergartens and attained BMIz and no evidence of effect modification by kindergarten type (P=0.571) (**Figure S2**).

## DISCUSSION

In a basic model with information on outcome, kindergarten type and birth weight, we found a lower risk of overweight after school entry among children in outdoor kindergartens compared to conventional kindergartens. However, when adding sociodemographic factors to the models, we found no differences in attained BMIz or risk of overweight between the two types of kindergartens. Likewise, we found no evidence that differences were modified by maternal education.

We have previously shown that children are more physically active in outdoor kindergartens compared to conventional kindergartens [18]. While the present study cannot confirm or refute causal effects, our results do suggest that potential effects of outdoor kindergartens on children’s activity levels or other potential mediators of body weight regulation are insufficient to prevent overweight or obesity at school entry.

Like Denmark, outdoor kindergartens are also part of the Swedish day-care system. Based on cross sectional data, Söderström and colleagues have previously found that Swedish children attending kindergartens with a high-quality outdoor environment less frequently had overweight than those attending kindergartens with a low-quality outdoor environment [14]. However, due to the cross-sectional design and the lack of adjustment for potentially important covariates, including parental BMI and socioeconomic status, the causal nature of this finding is highly uncertain. We found no previous studies directly comparing measures of attained adiposity in school aged children who had attended outdoor kindergartens and conventional kindergartens. However, as pointed out in a recent systematic review on nature and child health by Fyfe-Johnson and colleagues from 2021, only one-third of the published studies (n=45) found a lower risk of overweight among children with higher levels of nature exposure [11]. In our study, we found that children attending outdoor kindergartens had a lower risk of attaining obesity at school entry than children in conventional kindergartens before adjustment for important covariates such as familial BMI and socio-economic status. Indeed, the adjustment completely attenuated the observed associations. This further suggests that insufficient adjustment for confounding may have biased many of the previously published studies with positive results [11].

We also examined effect modification by maternal education, as low socioeconomic status is a strong predictor of childhood obesity [21] and previous results from the ODIN project have shown parental education to be higher among children attending outdoor kindergartens [22]. Although not specifically focused on obesity, there is also some evidence suggesting that children in low socioeconomic areas may have the greatest psychological benefits from exposure to high quality outdoor environments [23]. We found no evidence of effect modification in our analysis, but this could potentially also be due to a relatively high socioeconomic equality in the Danish population.

Our study has several strengths, including a prospective design with objectively measured information on study outcomes. We also had a relatively large sample size, providing us with sufficient statistical power to detect even small between-group difference in BMIz and risk of overweight. In addition, we had information on several covariates from the unique Danish health registers, including child and parental sociodemographic factors, allowing us to adjust for potential confounders.

On the other hand, our study also has some limitations. We had no information available on BMIz at kindergarten enrolment, which increases the risk of reverse causality. However, since we found no evidence of between group differences in the adjusted models, including the sensitivity analysis with added information on the last available measure of BMIz prior to kindergarten enrolment, it seems unlikely that adding a baseline measure of outcome to the model would have had a substantial impact on our results. We have previously shown a high degree of selection related to parental choice of kindergarten [22], and thus we can also not exclude that some unmeasured or residual confounding has remained. We had no information available on body composition, and we can therefore not rule out relevant between-group difference in attained fat or lean mass. Finally, our results originate from children attending kindergartens in two large Danish municipalities and may not be generalizable to children from other areas and we had a relatively large proportion of missing data. However, we found almost identical results when using multiple imputation to deal with missing data. Moreover, as we found no evidence of effect modification by maternal education or sex of the child it seems unlikely that results would be vastly different in Danish populations with other sociodemographic distributions.

In conclusion, our results do not support outdoor kindergartens as a potential intervention for healthy weight development. Given the observational design of our study, our results should be interpreted with caution, and randomized trials are warranted to confirm our findings.

## Data Availability

Data is stored on a secured server at Statistics Denmark (https://www.dst.dk/en/). Due to Danish legislation, the individual‐level data used in the present study cannot be made available.

