## Supplemental material for "Attained body mass index among children attending outdoor or conventional kindergartens"

### **Supporting figures**

#### **Figure S1.** Flowchart illustrating inclusion/exclusion of individuals from the Odin study.

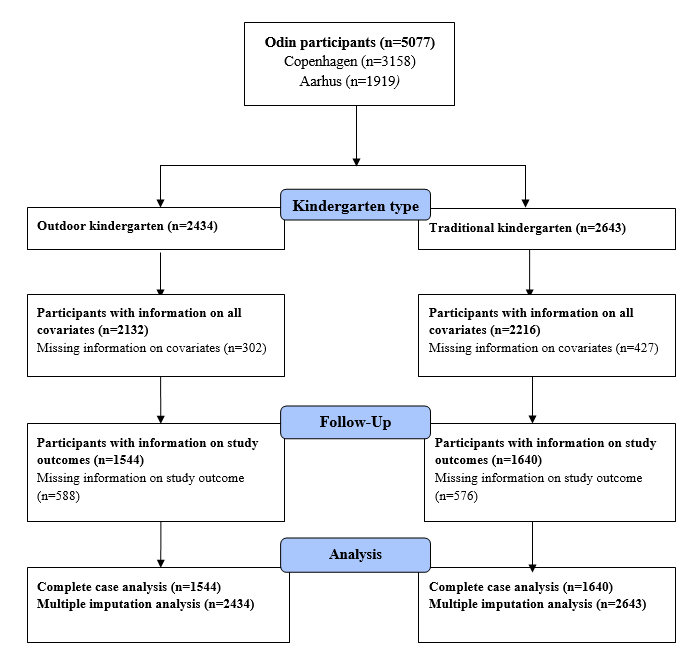

#### **Figure S2.** Association between years in kindergarten and attained BMI z-scores among children in outdoor

#### kindergartens and traditional kindergartens.

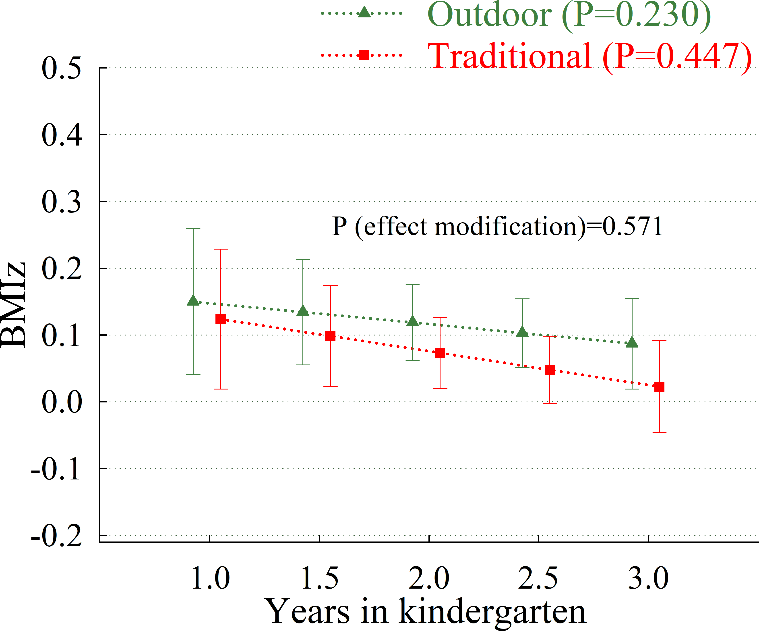

Abbreviations: BMIz, body mass index Z-score.

Results from model with information on outcome, total number of years spent in kindergarten, birth weight, maternal country of origin, maternal smoking during pregnancy, maternal pre-pregnancy body mass index, preterm birth, age at kindergarten enrolment.

### **Supporting tables**

#### **Table S1.** Attained BMI z-scores and risk of overweight among children in outdoor kindergartens versus traditional kindergartens adjusted for last recorded measure of BMI z-score prior to kindergarten enrolment (n=2681).

|  | Estimated difference | |
| --- | --- | --- |
|  | Difference or Odds Ratios (95% CI) | P-value |
| **BMIz** |  |  |
| Basic model ^1^ | -0.08 (-0.16, -0.01) | 0.043 |
| Full model ^2^ | 0.03 (-0.04, 0.11) | 0.388 |
| **Overweight** |  |  |
| Basic model | OR: 0.75 (0.62, 0.92) | 0.005 |
| Full model | OR: 0.92 (0.74, 1.12) | 0.481 |

Abbreviations: OR, Odds ratio; BMIz, body mass index Z-score.

^1^ Model with information on outcome, kindergarten type and birth weight.

^2^ Added information paternal education, paternal country of origin, maternal smoking during pregnancy, maternal pre-pregnancy body mass index, born preterm, age at kindergarten enrolment, total number of years spent in kindergarten and last recorded measure of BMIz prior to kindergarten enrolment.

#### **Table S2.** Attained BMI z-scores and risk among children in outdoor kindergartens versus traditional kindergartens with added information on paternal education and country of origin (n=2992).

|  | Estimated difference | |
| --- | --- | --- |
|  | Difference or Odds Ratios (95% CI) | P-value |
| **BMIz** |  |  |
| Basic model ^1^ | -0.08 (-0.15, -0.01) | 0.035 |
| Full model ^2^ | 0.03 (-0.04, 0.11) | 0.387 |
| **Overweight** |  |  |
| Basic model | OR: 0.77 (0.64, 0.93) | 0.008 |
| Full model | OR: 0.94 (0.77, 1.16) | 0.589 |

Abbreviations: OR, Odds ratio; BMIz, body mass index Z-score.

^1^ Model with information on outcome, kindergarten type and birth weight.

^2^ Added information maternal and paternal education, maternal and paternal country of origin, maternal smoking during pregnancy, maternal pre-pregnancy body mass index, born preterm, age at kindergarten enrolment, and total number of years spent in kindergarten.

#### **Table S3.** Attained BMI z-scores and risk of overweight among children in outdoor kindergartens versus traditional kindergartens in full sample (n=5,077). Missing information handled through multiple imputation.

|  | Estimated difference | |
| --- | --- | --- |
|  | Difference or Odds Ratios (95% CI) | P-value |
| **BMIz** |  |  |
| Basic model ^1^ | -0.08 (-0.15, -0.02) | 0.014 |
| Full model ^2^ | 0.05 (-0.02, 0.11) | 0.205 |
| **Overweight** |  |  |
| Basic model | OR: 0.79 (0.66, 0.93) | 0.005 |
| Full model | OR: 0.99 (0.82, 1.20) | 0.874 |

Abbreviations: OR, Odds ratio; BMIz, body mass index Z-score.

^1^ Model with information on outcome, kindergarten type and birth weight.

^2^ Added information maternal education, maternal country of origin, maternal smoking during pregnancy, maternal pre-pregnancy body mass index, born preterm, age at kindergarten enrolment, and total number of years spent in kindergarten.

### **Supporting text**

#### **Text S1.** Statistical analysis plan.

STUDY TITLE

Differences in attained body mass index and risk of overweight after school entry among children attending outdoor kindergartens or traditional kindergartens.

SUGGESTED AUTHORS

Sofus C. Larsen, Jeanett F. Rohde, Nanna J. Olsen, Jane N. Østergaard, Berit L. Heitmann, Ina O. Specht.

PURPOSE

In this statistical analysis plan, we describe the background and rationale for our study, in addition to general aspects of data preparation, sample size considerations and data analysis. The plan was set up and circulated with all authors prior to conducting any statistical analyses.

BACKGROUND

Despite increased awareness, the global prevalence of obesity among children and adolescents has increased from 0.9% to 7.8% among boys and from 0.7% to 5.6% among girls between 1975 and 2016 [1]. This has raised serious public health concerns, as childhood obesity is associated with both short- and long-term health consequences, including negative self-evaluation, bullying, social stigma, poor academic performance, symptoms of depression or anxiety [2, 3], numerous physical comorbidities [4] and an increased risk of mortality both in early [5] and later adulthood [6].

Once childhood obesity is established it is difficult to reverse [7] and obese children often remain obese in adulthood [8], strengthening the case for early intervention. However, the effect of existing prevention interventions are modest at best [9], indicating that alternative approaches may be needed to effectively prevent childhood overweight and obesity.

One of the many suggested explanations for the increased prevalence of childhood obesity is a decline in children’s outdoor activities. In support of this, cross-sectional studies have shown outdoor play time to be inversely associated with childhood body mass index (BMI), waist circumference and risk of obesity [10-12]. Moreover, Armstrong and colleagues (2015) have shown a higher density of parks or other recreational locations to be associated with a decrease in BMI z-score (BMIz) over time among 93 overweight children involved in a family-based weight management intervention [13].

Even though many of the published studies suggest a beneficial effect of outdoor activities on adiposity among children, the evidence is far from consistent [10]. In addition, there is a need to develop and evaluate interventions that can be implemented at a larger scale and in a cost–effective manner.

An ideal setting for conducting childhood interventions is the kindergarten, where many children spend a large part of their daily lives and establish some of the habits and health behaviours that may track into later childhood and adulthood. Major cities in Denmark and other Scandinavian countries have two different types of kindergartens: 1) a traditional kindergarten were the children have access to playgrounds but spend much of their day indoor [14], and 2) an outdoor kindergarten where most or all of the day is spent playing outside [14]. However, the effects of outdoor kindergartens on BMI or risk of obesity have not been thoroughly investigated.

Thus, combined with the unique Danish health registers and systematic measurements of height and weight performed by school health nurses, we have a unique opportunity to examine outdoor kindergartens as a potential intervention for preventing development of obesity later in childhood.

OBJECTIVE

Our primary objective is to examine whether children in outdoor kindergartens have attained a lower BMIz and risk of overweight after school entrance compared to children in traditional kindergartens, based on the first available measure of outcome recorded between 6 and 8 years of age.

Secondary objectives:

- Examine whether potential differences between kindergarten types are modified by socioeconomic status.
- Explore trajectories of attained BMIz and risk of overweight at 6, 7 and 8 years of age.

MATERIALS AND METHODS

**Study design**

As part of the *‘Outdoor kindergartens - the healthier choice?’* (ODIN) study, we have data on a total of 5077 children, of which 2434 attended outdoor kindergartens and 2643 attended traditional kindergartens. For the present study, we will include all children with information on selected covariates and at least one measure of BMIz after school entry (6 to 8 years of age).

**Outcomes**

Height and weight were measured by school health nurses during the first year of school and to some degree also during the subsequent years. From this information BMI has been calculated as weight in kilograms per height in meters squared. We have generated BMIz using a power transformation in increments of 0.1 years, applying national reference scores to the study population [15].

Primary outcome: Attained BMIz after school entrance.

Secondary outcome: Risk of attained overweight (including obesity) after school entrance [defined as a BMIz>1 (yes/no)]

**Covariates**

- Age at kindergarten enrolment (years).
- Time spent in kindergarten will be included as a continuous variable (years).
- Birth weight will be included as a continuous variable (g).
- Born preterm (yes/no).
- Maternal age (years).
- Maternal pre-pregnancy BMI.
- Maternal education categorised as basic, short, medium, and long. Basic (basic school 8th–10th class); Short (general upper–secondary education, short-cycle higher education or vocational education and training); Medium (medium-cycle higher education or bachelor); and Long (long-cycle education and PhD).
- Maternal country of origin categorised as Western and Non-Western.
- Maternal smoking during pregnancy (yes/no)*.*

*Additional covariates only included in sensitivity analyses*

- Paternal education categorised as basic, short, medium, and long.
- Paternal country of origin categorised as Western and Non-Western.
- Last available measure of BMIz prior to kindergarten enrolment (available on some of the children during the child’s first year of life measured by health nurses at the home of the child).

**Power calculation**

Assuming at least 3200 children have sufficient information to be included in the study, we will have more than 80% power to detect between group differences in BMIz of 0.1 or greater. Moreover, with 96% certainty the 95% CI will be no wider than 0.142 if we have data on 3200 children. The expected power and precision related to our primary outcome for a range of different sample sizes can be seen in **Figure 1.**

**Figure 1**. Estimated power (A) and estimated width of 95% CIs (B) for between group difference in our primary outcome (BMI z-scores) for a range of different sample sizes

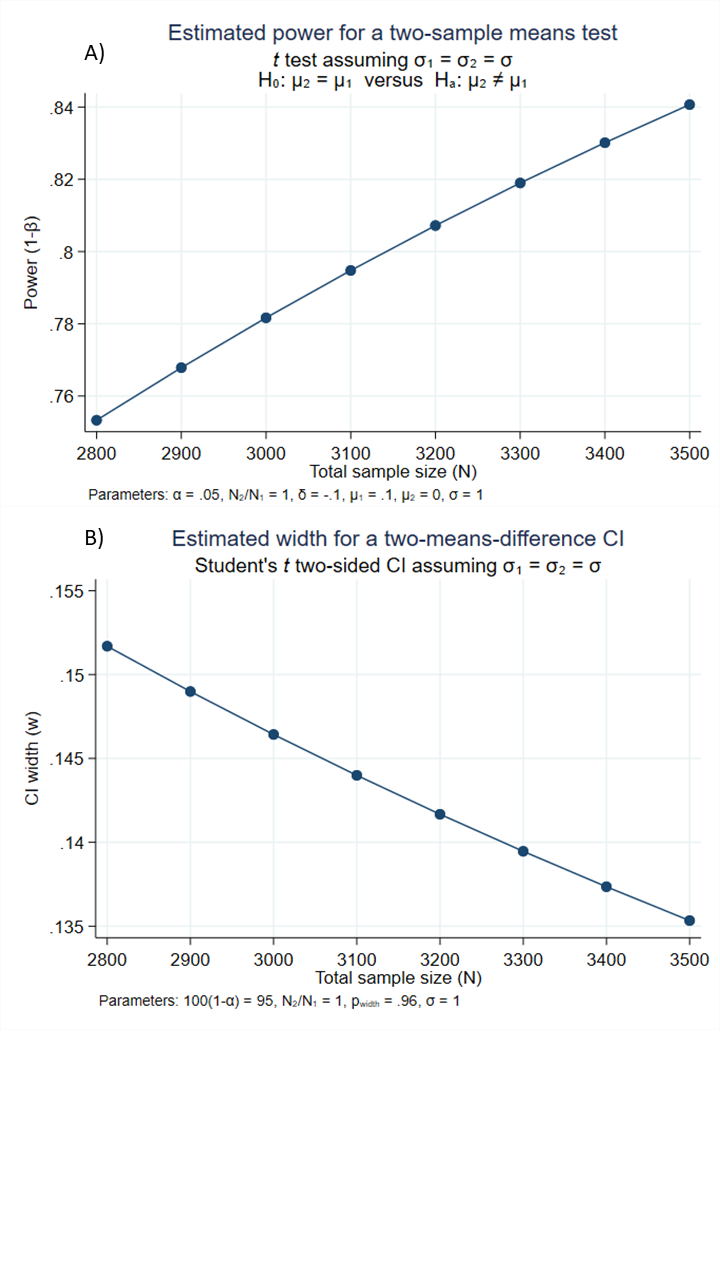

**Statistical analysis**

*Primary analyses*

Linear regression will be used to examine the mean difference in BMIz between the two groups based on the first recorded measure after school entry. We will present results from a basic model with information on BMIz, kindergarten type and birth weight only, and from a fully adjusted model with added information on additional covariates described above.

Using the same adjustment strategy, we will conduct logistic regression to examine the risk of attained overweight among children in outdoor kindergartens compared to traditional kindergartens based on the first recorded measure after school entry. It is well known that many people misinterpret odds ratios as risk ratios [16]. Thus, the postestimation command *adjrr* in STATA will be performed to attain adjusted estimates of absolute risks in addition to adjusted risk differences (ARD) and adjusted risk ratios (ARR) with corresponding 95% confidence intervals (95% CI) [16].

*Subgroup effects according to socioeconomic status*

Low socioeconomic status (SES) is a strong predictor of childhood obesity [17], and preliminary analyses from the ODIN project have shown parental education to be higher in outdoor kindergartens. Thus, in addition to adjustment for maternal education, we will conduct subgroup analyses specifically for children of low SES mothers (defined as mothers with a basic or short education) and high SES mothers (defined as mothers with a medium or long education). In addition, effect modification by SES will be tested by adding the new SES variable and a product term (*kindergarten type* *× SES*) to the fully adjusted models after removing the original maternal education variable from the model.

*Trajectories of attained BMIz and overweight*

The same overall strategy will be used to explore trajectories of attained BMIz and overweight at specific ages after school entry (i.e. 6, 7 and 8 years of age). These analyses will be restricted to children with at least one measurement of BMIz at 6 years of age. A last observation carried forward approach will then be used for children with missing information at 7 and/or 8 years of age.

All statistical tests will be two-sided with a significance level at 0.05. All statistical analyses will be performed using Stata/SE 16 (StataCorp LP, College Station, Texas, USA). Figures will be produced using SigmaPlot 13.0 (San Jose, CA, USA).

*Sensitivity analyses*

We do not have a baseline measure of outcome available. However, in addition to birth weight, most of the children had weight and length measured during the first year of life by health nurses at the home of the child. Thus, in a sensitivity analysis we will add the last available measure of BMIz prior to kindergarten enrolment as a covariate.

For the primary analyses, we will use maternal education as a proxy for the child's SES. Similarly, we will use maternal country of origin as a proxy for the geographical origin of the child. However, sensitivity analyses will be conducted adding paternal information to the models.

Since BMIz takes into account differential BMI trajectories by sex and age, we will not include sex as potential confounders. However, effect modification by sex will be tested by adding the variable *sex* and a product term (*kindergarten type* *× sex*) to the fully adjusted models. Subgroup analyses will only be conducted if statistically significant interactions are identified.

In the primary analyses we will only include participants with complete information on all covariates, but as a sensitivity analysis multiple imputation will be implemented for all covariates with missing values.

Finally, we will examine a potential dose–response relationship using information on the total time spend in outdoor kindergarten (continuous variable) for each child.

RESULTS

This section presents the templates of figures and tables related to statistical analyses planed a priori. Some additional sensitivity analyses may be added subsequently.

**Table 1.** Participant characteristics of children in outdoor kindergartens versus traditional kindergartens. Results presented as mean (SD) unless otherwise stated.

|  | Outdoor kindergarten (n=1544) | Traditional kindergarten (n=1640) |  |
| --- | --- | --- | --- |
|  | Mean (SD) | Mean (SD) | P-value |
| Female sex, n [%] |  |  |  |
| Age at kindergarten enrolment, years |  |  |  |
| Age at first school measurement, years |  |  |  |
| Time spent in kindergarten, years |  |  |  |
| Birth weight, g |  |  |  |
| Born preterm, n [%] |  |  |  |
| Maternal age, years |  |  |  |
| Maternal pre-pregnancy body mass index |  |  |  |
| Maternal education ^1^ |  |  |  |
| Basic, n [%] |  |  |  |
| Short, n [%] |  |  |  |
| Medium, n [%] |  |  |  |
| Long, n [%] |  |  |  |
| Maternal country of origin |  |  |  |
| Non-Western, n (%) |  |  |  |
| Mother smoked during pregnancy, n (%) |  |  |  |

*Abbreviations: BMIz, body mass index Z-score*

**Table 2.** Attained BMI z-scores and risk of overweight among children in outdoor kindergartens versus traditional kindergartens.

|  | Mean or % of participants (SE) | | Estimated difference | |
| --- | --- | --- | --- | --- |
|  | Outdoor kindergarten | Traditional kindergarten | Difference, ARD or ARR (95% CI) | P-value |
| **BMIz** |  |  |  |  |
| Basic model ^1^ |  |  |  |  |
| Full model ^2^ |  |  |  |  |
| **Overweight** |  |  |  |  |
| Basic model |  |  | ARD  ARR |  |
| Full model |  |  | ARD  ARR |  |

*Abbreviations: ARD, adjusted risk difference; ARR, adjusted risk ratio; BMIz, body mass index Z-score.*

*^1^ Model with information on outcome, kindergarten type and birth weight.*

*^2^ Added information maternal education, maternal country of origin, maternal smoking during pregnancy, maternal pre-pregnancy body mass index, born preterm, age at kindergarten enrolment, and total number of years spent in kindergarten.*

**Figure 2 (template).** Hypothetical attained BMI z-scores (A) and risk of overweight (B) among children in outdoor kindergartens versus traditional kindergartens. Results stratified by maternal socioeconomic status.

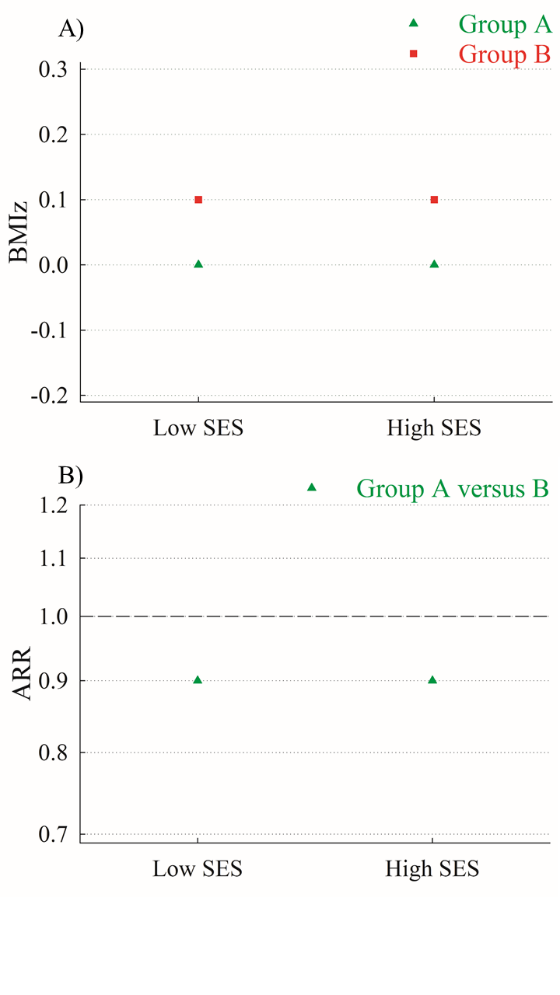

*Abbreviations: ARR, adjusted risk ratios; BMIz, body mass index Z-score; SES, socioeconomic status.*

*Low SES is defined as mothers with a basic or low education. High SES is defined as mothers with a medium or high education.*

*Results from model with information on outcome, kindergarten type, birth weight, maternal country of origin, maternal smoking during pregnancy, maternal pre-pregnancy body mass index, preterm birth, age at kindergarten enrolment, and total number of years spent in kindergarten.*

**Figure 3 (template).** Hypothetical trajectories of attained BMIz (A) or overweight (B) at 6, 7 and 8 years of age.

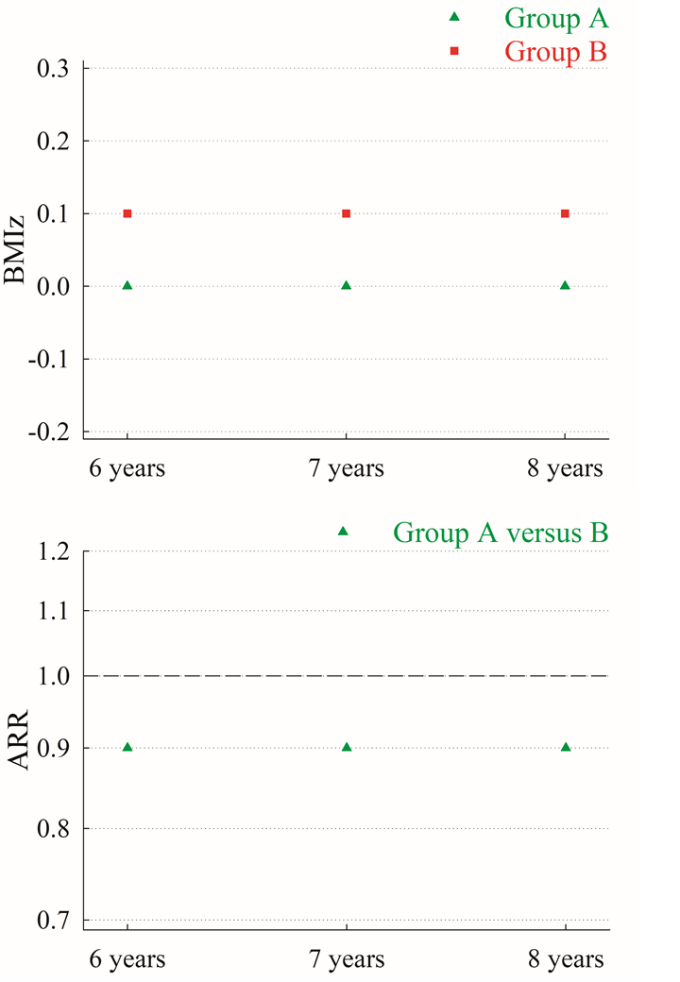

*Abbreviations: BMIz, body mass index Z-score; ARR, adjusted risk ratios*

*Results from model with information on outcome, kindergarten type, birth weight, maternal country of origin, maternal smoking during pregnancy, maternal pre-pregnancy body mass index, preterm birth, age at kindergarten enrolment, and total number of years spent in kindergarten.*

14.	MN Christoffersen, Højen-Sørensen AKH, Laugesen L. Danish title: Daginstitutionens betydning for børns udvikling – en forskningsoversigt. English translation: The significance of the day care institution for children's development - a research overview. SFI Det nationale Forskningscenter for velfærd. 2014.

15.	Nysom K, Mølgaard C, Hutchings B, Michaelsen KF. Body mass index of 0 to 45-y-old Danes: reference values and comparison with published European reference values. Int J Obes Relat Metab Disord. 2001;25(2):177-84. Epub 2001/06/19. doi: 10.1038/sj.ijo.0801515. PubMed PMID: 11410817.

16.	Norton EC, Miller MM, Kleinman LC. Computing adjusted risk ratios and risk differences in Stata. Stata Journal. 2013;13(3):492-509.

17.	Frederick CB, Snellman K, Putnam RD. Increasing socioeconomic disparities in adolescent obesity. Proc Natl Acad Sci U S A. 2014;111(4):1338-42. Epub 2014/01/30. doi: 10.1073/pnas.1321355110. PubMed PMID: 24474757; PubMed Central PMCID: PMCPMC3910644.
